# A standard coil placement for reliable transcranial magnetic stimulation of the frontoparietal depression network: the ‘F5-AF7 method‘

**DOI:** 10.64898/2026.01.25.26344778

**Authors:** Maximilian Lueckel, Kathrin Kachel, Jan Engelmann, Til Ole Bergmann, Florian Müller-Dahlhaus

**Author notes:** Contributed equally (Til Ole Bergmann, Florian Müller-Dahlhaus). Corresponding authors: Maximilian Lueckel, Florian Müller-Dahlhaus.

## Abstract

**Background:** The frontoparietal network (FPN) has been strongly implicated in both the development of and recovery from major depressive disorder (MDD), making it a promising target for transcranial magnetic stimulation (TMS) therapy of MDD. However, commonly used TMS targeting approaches often co-stimulate multiple neighboring functional brain networks to varying degrees across individuals. We therefore aimed to develop a clinically feasible, standardized TMS coil placement that enables stronger and more selective stimulation of the FPN without requiring individual neuroimaging or neuronavigation.

**Methods:** We optimized the placement of a prototypical figure-of-eight coil in a population-based brain template by maximizing simulated electric field (E-field) strength within a representative FPN cluster. Based on this optimized placement, we derived a practical heuristic using EEG electrode positions and simple scalp measurements. The FPN-optimized placement and its heuristic were validated by comparing E-field hotspot coverage of the FPN and other functional networks against a clinically established, standard coil placement (Beam F3 method) in precisely mapped brains of 20 healthy individuals (15 female) and 20 patients with MDD (7 female). We further assessed robustness of FPN stimulation to coil tilt inaccuracies and the role of coil orientation.

**Results:** The optimized heuristic places the center of the coil over the F5 electrode and orients its handle along the F5-AF7 axis. This placement yielded significantly stronger and more selective FPN coverage than the established clinical approach. Targeting was largely robust to tilt inaccuracies but sensitive to rotational deviations.

**Discussion:** This scalp-based F5-AF7 TMS coil placement enables selective and reliable targeting of the frontoparietal depression network in routine clinical settings. Whether it improves antidepressant efficacy compared to established targeting strategies should be evaluated in future clinical trials.

## 1 Introduction

Repetitive transcranial magnetic stimulation (rTMS) is an established treatment for major depression (1–3). However, response and remission rates to standard clinical rTMS protocols are modest and variable (4,5). One suggested factor contributing to this variability is the exact placement of the TMS coil (6), which determines the specific functional brain networks engaged by the stimulation (7–9). Accordingly, treatment effectiveness of rTMS has been related (mostly retrospectively) to the individual functional network connectivity (FC) of the stimulated cortical area, seemingly resulting in better treatment outcomes when applied closer to the person-specific spot in the left dorsolateral prefrontal cortex (DLPFC) whose resting-state activity is most anticorrelated to that of the subgenual anterior cingulate cortex (sgACC) ((10–16); but see (17)).

Following this finding, protocols have been developed that try to prospectively personalize TMS coil placement based on individually acquired functional magnetic resonance imaging (MRI) data and derived connectivity measures (18–22). Such connectivity-guided approaches are sometimes additionally combined with individual simulations of the TMS-induced electric field (E-field) to further optimize targeting precision and specificity (7,23,8,24,9). While these personalized, MRI-guided targeting strategies are conceptually appealing and promise to increase the clinical effectiveness of TMS (18,20,21,19), it is currently unclear whether or when they will be applicable, affordable, and scalable in clinical practice (25) and replace existing fast, cheap, and widely used heuristics for TMS coil placement, such as the Beam F3 method (which uses simple scalp measurements to place the TMS coil on the estimated F3 position of the International 10-20 system for electroencephalography (EEG) electrodes; (26)).

Another critical question pertains to the specific functional networks targeted by TMS. The typical stimulation area in TMS depression treatment, i.e., the (left) DLPFC, comprises multiple differentiable functional networks (27,28). Next to nodes of the cingulo-opercular or action-mode network (CON/AMN; (29)) and salience network (SN; (30,31)), the DLPFC contains large representations of the frontoparietal network (FPN; (32)) (Figure 1A). The FPN has not only been broadly related to cognitive/executive control, emotion regulation, and working memory (32–36) – key functions often impaired in depression – but more importantly, it has been causally implicated in the development of and relief from depression: Padmanabhan et al. (37) found that locations of brain lesions associated with depression were functionally connected to a network strongly resembling the FPN (Figure 1A). This finding was further supported and extended by Siddiqi and colleagues (38), showing that not only lesions, but also target sites of TMS and deep brain stimulation that modulate symptoms of depression exhibited strong functional connectivity to a similar frontoparietal circuit. Together, these findings suggest a common frontoparietal depression network and, thus, pose the FPN as a promising target network for TMS depression treatment.

**Figure 1.**
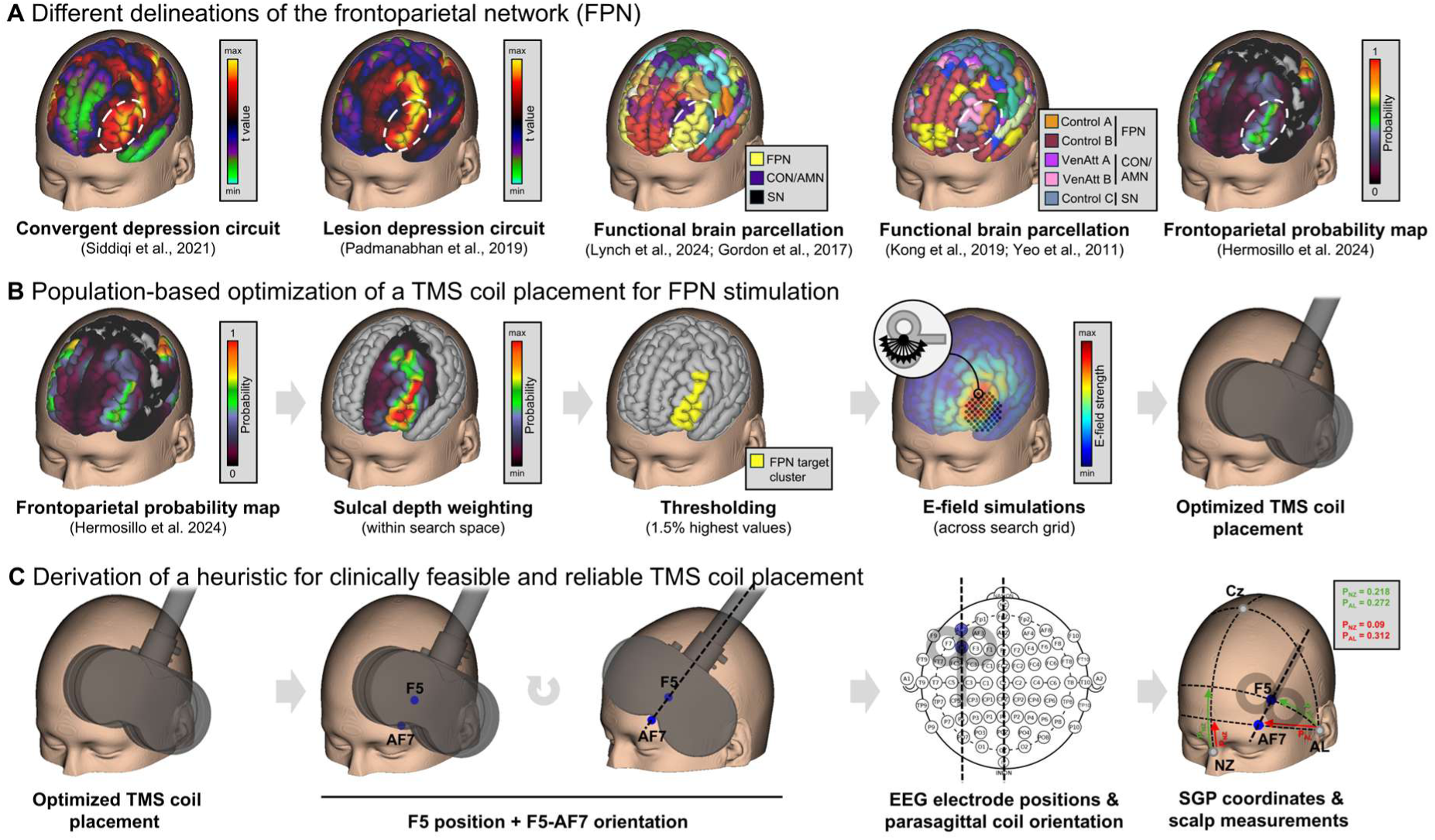
Derivation of an optimized TMS coil placement and a clinical heuristic for targeting the FPN. **A**: Illustration of different delineations of the FPN. **B**: Overview of the procedure for optimizing the TMS coil placement for FPN stimulation using simulations of the TMS-induced E-field in a search grid (in MNI standard space). **C**: Derivation of a clinical heuristic for reliably reproducing the population-based FPN-optimized TMS coil placement based on EEG electrode positions and SGP coordinate system-based scalp measurements.

Yet, the considerable interindividual variability in functional network topographies (27,28) as well as the non-uniform spread of the TMS-induced E-field across the cortex (39,40) make it difficult to selectively target a specific network with TMS (9). Combined precision mapping of individual functional network boundaries and E-field simulations have shown that both clinically used heuristic and personalized (sgACC-)FC-guided TMS of the DLPFC is often highly unspecific: both targeting strategies appear to primarily co-stimulate the CON/AMN, SN, and FPN to different degrees in different individuals (cf. Figure 2; (8,7)). However, given the proposed causal role of the FPN in depression, a TMS coil placement that more strongly and selectively targets and engages the FPN, while less strongly stimulating neighboring off-target networks, might be desirable and potentially improve clinical effectiveness of TMS depression treatment.

**Figure 2.**
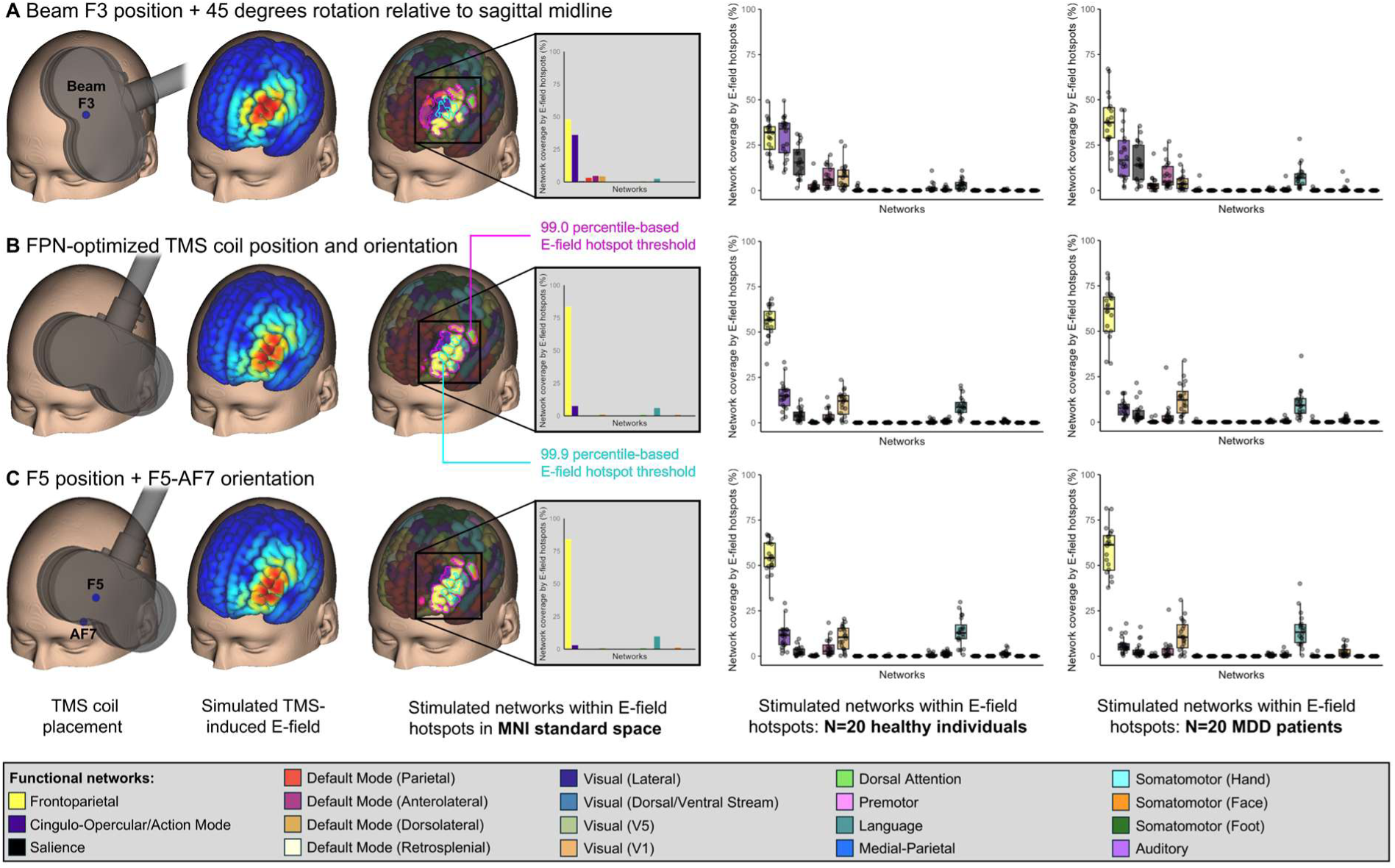
Percent coverage of functional networks (Lynch et al. (31) parcellation) by simulated E-field hotspots. Functional network coverage resulting from (**A**) a standard clinical TMS coil placement (i.e., TMS coil centered on the Beam F3 position and oriented 45 degrees relative to the sagittal midline), (**B**) the population-based FPN-optimized TMS coil placement, and (**C**) its F5-AF7 EEG electrode-based heuristic (i.e., TMS coil centered on the F5 EEG electrode position with its handle oriented along the F5-AF7 axis) in MNI standard space, healthy individuals (N=20), and MDD patients (N=20). Note that the depicted values represent the average over ten different percentile-based E-field thresholds (see Methods and Materials for more details). Exemplary E-field hotspots are illustrated for the lowest (99.0 percentile; magenta outline) and highest (99.9 percentile; cyan outline) threshold.

We therefore aimed to determine an optimized, but generally applicable and standardized TMS coil placement (i.e., position and orientation) for more selective and reliable targeting of the depression-relevant FPN, utilizing recently developed methods from connectivity- and E-field-guided precision neuromodulation (7,41). Importantly, we additionally aimed to provide a clinically practical heuristic to easily but reliably and precisely determine this FPN-optimized coil placement in any given patient, without the need for individual functional and/or anatomical MRI data and neuronavigation.

## 2 Methods and Materials

### 2.1 Definition of the stimulation target

We first quantitatively confirmed that the causal depression network maps by Siddiqi et al. (38) and Padmanabhan et al. (37), particularly their DLPFC-portion (encircled in Figure 1A), align with common delineations of the FPN (Figure 1A). More specifically, using the Dice coefficient to quantify spatial network overlap and spin test permutations to determine its statistical significance (as implemented in the Network Correspondence Toolbox (42) in Python 3.11), we compared (surface-mapped and thresholded versions of) the two depression circuit maps (Figure S1) against discrete functional network parcellations by i) Gordon et al. (27) (17 networks; Figure S1), ii) Lynch et al. (31) (a refined version of the Gordon parcellation with 19 networks; Figures 1A and S1), iii) Yeo et al. (43) (17 networks; Figure S1), and iv) Kong et al. (44) (a refined version of the Yeo parcellation with 17 networks; Figures 1A and S1), as well as v) probabilistic maps of 14 commonly observed large-scale functional networks (Figure 1A), provided as part of a precision functional brain atlas by Hermosillo et al. (45). All maps were available in fs_LR_32k surface space (46). Consistently across all reference networks and parcellations, the two causal depression network maps significantly overlapped with the FPN (Dice coefficient range: 0.3183 – 0.5178; all p’s < .001; for details and overlap with other networks, see Figure S1 in the Supplement).

Next, we selected the probabilistic FPN map of the precision functional brain atlas (45) as a representative standard template for the average, population-based topography of the FPN (Figure 1B). We then restricted this map to the extended stimulation area of interest, i.e., the left frontal cortex (including the left DLPFC), and weighted the remaining vertex values by multiplying them with the corresponding sulcal depth value of each vertex (Figure 1B). Thereby, the probability weight of areas within sulci was decreased, while upweighting the relative importance of areas on the gyral crown, where the TMS-induced E-field has its strongest effect (39,47). This masked and weighted map was then thresholded at the 1.5% highest values to define a coherent, representative FPN target cluster concentrated on gyral crowns within the left DLPFC (Figure 1B).

### 2.2 Electric field simulations and optimization of TMS coil placement in standard space

To construct a realistic biophysical head and brain model for simulations of the TMS-induced E-field in MNI standard space, we first subjected the MNI152NLin2009cAsym T1- and T2-weighted template images (48) to the standard Freesurfer recon-all pipeline (49). Afterwards, the SimNIBS charm pipeline was run on the same anatomical input images (SimNIBS version 4.5; (50,51)), including the surface files generated by Freesurfer as additional inputs, to achieve higher accuracy of charm’s grey and white matter segmentations and surface reconstructions.

Next, simulations of the TMS-induced E-field were performed using SimNIBS utilities (SimNIBS version 4.5; (51)) and a model of the MagVenture Cool-B65 coil (52). E-field simulations were run in a search grid of 828 theoretical TMS coil placements (i.e., 69 coil positions, 5 mm apart from each other, and 12 coil rotations per position, covering 180 degrees in steps of 15 degrees, with a 1 mm coil-scalp-distance) that was defined on the reconstructed scalp of the MNI head, centered around the point on the scalp closest to the geometric center of the FPN target cluster (MNI coordinates: X = - 45.20, Y = 56.02, Z = 45.21) (Figure 1B). We used the minimum possible stimulation intensity of dI/dt = 1 A/µs for all simulations, given that the E-field varies linearly with dI/dt and its relative spatial distribution is therefore independent from the absolute stimulation intensity (7).

Each resulting simulated E-field was interpolated to the reconstructed central grey matter surface and resampled to fs_LR_32k space (46). We then computed the sum of all vertex-wise products between each respective E-field map and the FPN target cluster map. Eventually, the TMS coil placement that maximized this sum was considered optimal for stimulating the left DLPFC-portion of the FPN (in MNI standard space) as it induced an E-field that maximally overlapped with the FPN target cluster (53). In the following, this TMS coil placement will be called the “population-based FPN-optimized” placement.

### 2.3 Derivation of a clinical heuristic

As the resulting population-based FPN-optimized TMS coil placement is defined by coordinates in MNI standard space, realizing this placement in a given individual would usually require some form of neuronavigation (based on either the MNI template or individual anatomical MRI data). However, neuronavigation systems are not always available in clinical settings, which may hinder the clinical translation of improved FPN targeting. Therefore, we aimed to infer a simpler way of determining the respective coil placement in routine clinical practice according to the following procedure.

We first identified the EEG electrode closest (i.e., with the shortest Euclidean distance) to the center of the population-based FPN-optimized TMS coil placement, using the EEG electrode positions automatically calculated by SimNIBS’ charm as part of head model generation (54,55). This approach was inspired by the common clinical practice of using EEG electrode positions on the scalp to define the position of the TMS coil center (e.g., F3 in the Beam F3 method; (26)).

To also provide specific recommendations for the optimal orientation of the TMS coil, which is often neglected but increasingly recognized as a crucial factor for TMS coil placement as it strongly influences how the E-field distributes across the cortex (e.g., (39,56)), we determined the angle that the optimal TMS coil placement deviated from an orientation parallel to the sagittal midline. Furthermore, to also derive a clinical heuristic for coil orientation, we determined the EEG electrode that, when drawing a straight line from the electrode position that heuristically defines the TMS coil center position (see previous paragraph), would best recreate the direction of the optimized coil handle orientation (cf. Figure 1C). In the following, this TMS coil placement will be called the “EEG electrode-based heuristic” placement.

Lastly, to provide a scalp measurement heuristic that is immediately applicable in clinical practice, we used the EEG2CPC mapping tool (https://transcranial-brain-atlas.org/eeg2cpc/) to derive the coordinates for both of the previously determined EEG electrode positions in scalp geometry-based parameter (SGP) space (57), which is based on the continuous proportional coordinate (CPC) standard scalp space (58,59). In short, SGP space enables definition of any point (and, thus, EEG electrode position) on an individual’s scalp in terms of two coordinates (P_NZ_ and P_AL_), based on distances between five individually measured head reference points (nasion (NZ), inion (IZ), left (AL) and right (AR) preauricular points, and the center point (Cz) of the NZ-IZ and AL-AR lines; for details, refer to (57)). Importantly, SGP-defined EEG electrode positions (particularly F3) have been shown to be more precise than other established scalp-measurement methods (e.g., Beam F3; (60,61)).

### 2.4 Validation of FPN stimulation in standard space and individual subject data

To confirm that the resulting population-based FPN-optimized TMS coil placement and its EEG electrode-based heuristic indeed result in stronger and more selective stimulation of the FPN, we compared them against a standard clinical TMS coil placement, with the coil center position defined by the Beam F3 method (26) and the coil handle oriented 45 degrees relative to the sagittal midline (e.g., (62–64)). To determine the scalp measurement-based Beam F3 location on a SimNIBS-derived head model, we used the following procedure: First, to identify the four anatomical landmarks required for performing the Beam F3 method on the head model (i.e., nasion, inion, left and right tragus), we used the respective coordinates automatically calculated by SimNIBS’ charm as part of head model generation; second, to measure the required distances along the mesh surface of the head model (i.e., nasion-to-inion, left-to-right tragus, and head circumference), we used Matlab-based tools provided by (61) and (65) (note that, following recommendations by (66) and (61), the Fpz and Oz electrode positions, as derived by SimNIBS’ charm, were used to calculate head circumference); third, these distances were then used to calculate the X and Y values according to the clinically used web version of the Beam F3 formula (https://www.clinicalresearcher.org/F3/calculate.php) and to identify the scalp coordinate that would define the center position of the TMS coil (note that, here, we used the modified Beam F3 method with an adjusted Y value according to (66)).

Further, for each of the three TMS coil placements of interest (i.e., population-based FPN-optimized, EEG electrode-based heuristic, and Beam F3 + 45 degrees orientation), we used SimNIBS (version 4.5; (51)) and the MagVenture Cool-B65 TMS coil model (52) to simulate the respective TMS-induced E-fields. The resulting E-field maps were interpolated to the reconstructed central grey matter surface, resampled to fs_LR_32k space (46), and thresholded at ten different percentile thresholds (99.0% to 99.9%, in 0.1% steps, in order to avoid potential bias from only selecting one specific threshold; (7)) to define ten E-field “hotspot” areas of differing spatial extent. Employing refined versions of two commonly used functional brain parcellations from Gordon et al. (27) (refined by (31); Figure 1A, Figure 2) and Yeo et al. (43) (refined by (44); Figure 1A, Figure S1 in the Supplement), we then extracted, for each of the ten hotspot definitions, the percentage of the total hotspot area covering each functional network as defined by the respective parcellation. These fractions of E-field hotspot coverage were eventually averaged across the ten hotspot thresholds, separately for each network, and statistically compared between the three TMS coil placements (note that, for statistical comparisons, we focused on coverage of the FPN, CON/AMN, and SN, i.e., the three most strongly represented functional networks within the (left) DLPFC, while E-field hotspot coverage of all functional networks is plotted in Figure 2 and Figure S3).

Importantly, this entire procedure was not only performed in MNI template data, but also in individual subject data. For that, we used anatomical and functional MRI data from N=20 healthy individuals (15 females) and N=20 patients with Major Depressive Disorder (MDD; 7 females; for sample characteristics, MRI sequence parameters, and data processing steps, refer to the Supplement; all study procedures were approved by the local ethics committee in accordance with the Declaration of Helsinki (ethics vote no. 2019-14452 and 2020-15333), and written informed consent was provided by each participant). Based on their resting-state fMRI scans, we precisely mapped each subject’s individual functional network topography using the Multi-Session Hierarchical Bayesian Modelling (MS-HBM) approach, which, in short, relies on group-level spatial priors derived from independent training data to estimate individual-specific network parcellations (for details, see (44)). This approach was implemented in Matlab R2022a, using priors of the functional networks defined by both the 19-network parcellation by Lynch et al. (31) (Figure 1A, Figure 2) and the 17-network parcellation by Kong et al. (44) (Figure 1A, Figure S3).

### 2.5 Robustness against inaccuracies in TMS coil placement and validation of the optimized coil orientation

In clinical practice, it is not always assured that the TMS coil is placed perfectly tangentially to the scalp. Rather, the TMS coil may be slightly tilted along the axis of the coil handle (y-axis; Figure 4 and S5) and/or the axis of the two coil windings (x-axis; in the case of a figure-of-eight coil; Figure 4 and S5) (cf. Figure 1 in (56), “roll” and “pitch”, respectively). This may change the topography of and, thus, the overlap between the TMS-induced E-field (hotspot) and targeted functional brain networks, resulting in certain networks being more or less strongly stimulated. Hence, to test the robustness of FPN targeting against slight inaccuracies in TMS coil tilt, we simulated the TMS-induced E-fields for four additional tilts (-10, -5, +5, +10 degrees) along the coil’s y- and x-axis for each individual, using the original heuristic EEG electrode-based TMS coil placement (with 0 degrees tilt) as reference. Relatedly, to also demonstrate that stronger and more selective FPN targeting not only depends on the proposed center position of the TMS coil, but is also influenced by its orientation, we simulated the TMS-induced E-fields for 11 additional coil orientations (-90 to +75 degrees, in 15 degree steps) relative to the original, optimized orientation of the proposed heuristic EEG electrode-based TMS coil placement (with 0 degrees rotation). In all cases, we again extracted the fraction of functional networks covered by the respective E-field hotspots (averaged across ten percentile-based hotspot thresholds from 99.0% to 99.9%, in steps of 0.1%, as described in more detail above).

### 2.6 Statistical analysis and visualization

We used repeated-measures ANOVAs and paired post-hoc t-tests in R (version 4.3.3; (67)) to compare the individually determined (average) fractions of the FPN, CON/AMN, and SN covered by hotspots of simulated TMS-induced E-fields (i) between the three targeting approaches (Beam F3 + 45 degrees orientation, population-based FPN-optimized, EEG electrode-based heuristic) as well as (ii) between the optimal 0 degree tilt and orientation condition and all additional tilt (-10, -5, +5, and +10 degrees along both the y- and x-axis of the TMS coil) and orientation (-90, -75, -60, -45, -30, -15, +15, +30, +45, +60, and +75 degrees along the z-axis of the TMS coil) conditions. False discovery rate was used for multiple comparisons correction, and the level for statistical significance was set to α = 0.05. Note that, independent of statistical significance, we only report and interpret effects of at least medium size (i.e., Cohen’s |d| ≥ 0.5), given that, due to the repeated measures design, even negligible to small effects tend to be (highly) significant, but are not practically relevant.

Bar, raincloud, and tile plots were created using the ggplot2 package (68) in R (version 4.3.3; (67)). The 3D-reconstructed MNI brain (pial surface) and the overlaid network maps and parcellations were visualized using Connectome Workbench viewer (69). 3D Slicer (version 5.4.0; (70)) was used to visualize the 3D-reconstructed MNI head surface and TMS coil model.

## 3 Results

### 3.1 Optimized TMS coil placement for FPN stimulation and its clinical EEG electrode-based heuristic

As illustrated in Figure 1B and quantified in Table 2, our optimization procedure resulted in a TMS coil placement that induced a (simulated) E-field which strongly overlaps with the representative FPN target cluster (in MNI standard space). The MNI coordinates of the TMS coil center on the scalp and the closest point in the brain are given in Table 1. The angle of the coil handle orientation relative to the sagittal midline was 1.1 degrees, i.e., the coil handle was essentially parallel to the sagittal midline.

**Table 1.** Overview of coordinates for the population-based FPN-optimized TMS coil placement and its clinical EEG electrode-based heuristic

|  | **Population-based FPN-optimized**<br>**TMS coil placement**<br>**[MNI coordinates]** | **Heuristic EEG electrode-based**<br>**TMS coil placement**<br>**[EEG electrode positions;**<br>**SGP coordinates]** |
| --- | --- | --- |
| **TMS coil center** | Scalp: X = -62.19, Y = 49.16, Z = 19.03<br>Brain: X = -50.68, Y = 41.58, Z = 15.61 | F5;<br>$P_{NZ} = 0.218$, $P_{AL} = 0.272$ |
| **TMS coil**<br>**orientation** | Parallel to sagittal midline | (Axis from F5 to) AF7;<br>$P_{NZ} = 0.09$, $P_{AL} = 0.312$ |

**Table 2.**
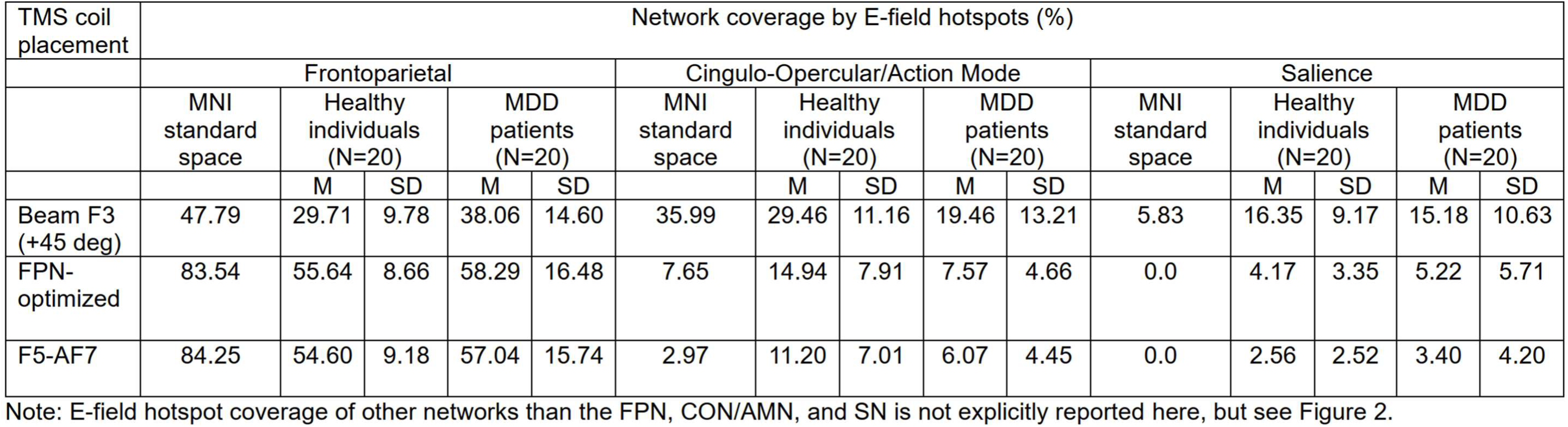
Functional network coverage by E-field hotspots of different TMS coil placements.

For the clinical heuristic, we found that the optimized TMS coil position was very close to the F5 EEG electrode position (Figure 1C), with an Euclidean distance of only 4.33 mm. Furthermore, the coil handle orientation defined by a line running from the F5 to the AF7 EEG electrode position almost perfectly reproduced the optimized coil handle orientation (Figure 1C), deviating by only 1.47 degrees. SGP coordinates for both EEG electrodes are given in Table 1 and Figure 1C.

### 3.2 Validation of the population-based FPN-optimized and heuristic F5-AF7 TMS coil placement

Figure 2 illustrates and Table 2 summarizes the (average) percent coverage of the FPN, CON/AMN, and SN by simulated E-field hotspots resulting from a standard clinical TMS coil placement (i.e., TMS coil centered on the Beam F3 position and oriented 45 degrees relative to the sagittal midline), the population-based FPN-optimized TMS coil placement, and its F5-AF7 EEG electrode-based heuristic (i.e., TMS coil centered on the F5 EEG electrode position with its handle oriented along the F5-AF7 axis) in MNI standard space, healthy individuals (N=20), and MDD patients (N=20).

Statistical comparisons between the three targeting approaches showed that, in comparison to the standard Beam F3-based TMS coil placement, E-field hotspots of the newly derived population-based FPN-optimized and heuristic F5-AF7 TMS coil placements covered (i) a significantly larger fraction of the FPN in both healthy individuals (p_FDR_ < .001, d = 2.79 and p_FDR_ < .001, d = 2.62, respectively; Figure 3A, left) and MDD patients (p_FDR_ < .001, d = 1.29 and p_FDR_ < .001, d = 1.25, respectively; Figure 3B, left), (ii) a significantly smaller fraction of the CON/AMN in both healthy individuals (p_FDR_ < .001, d = -1.41 and p_FDR_ < .001, d = -1.80, respectively; Figure 3A, middle) and MDD patients (p_FDR_ < .001, d = -0.85 and p_FDR_ < .001, d = -1.13, respectively; Figure 3B, middle), and (iii) a significantly smaller fraction of the SN in both healthy individuals (p_FDR_ < .001, d = -1.43 and p_FDR_ < .001, d = -1.79, respectively; Figure 3A, right) and MDD patients (p_FDR_ < .001, d = -1.10 and p_FDR_ < .001, d = -1.33, respectively; Figure 3B, right).

**Figure 3.**
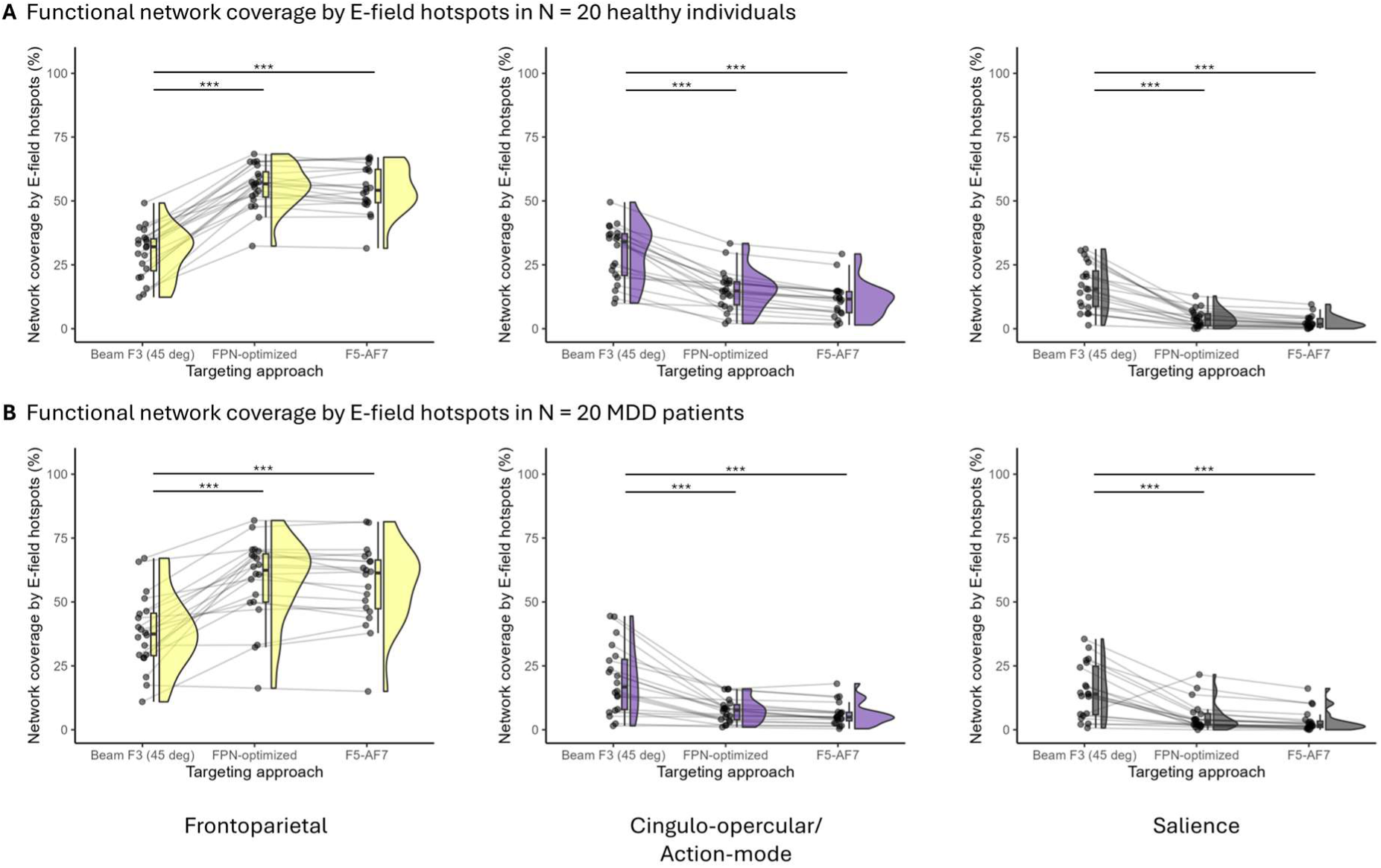
Percent coverage of functional networks (Lynch et al. (31) parcellation) by simulated E-field hotspots. Coverage of the frontoparietal network (FPN; left), cingulo-opercular/action mode network (CON/AMN; middle), and salience network (SN; right) by simulated E-field hotspots resulting from a standard clinical TMS coil placement (i.e., TMS coil centered on the Beam F3 position with its handle oriented 45 degrees relative to the sagittal midline), the population-based FPN-optimized TMS coil placement, and its F5-AF7 EEG electrode-based heuristic (i.e., TMS coil centered on the F5 EEG electrode position and oriented along the F5-AF7 axis) in (**A**) healthy individuals (N=20) and (**B**) MDD patients (N=20). Note that the depicted values represent the average over ten different percentile-based E-field thresholds (see Methods and Materials for more details). *** p_FDR_ < .001 and d ≥ 0.50.

While these results are based on the network parcellation by Lynch et al.(31), the same pattern of results was replicated using a different parcellation of functional brain networks by Kong et al. (44) (see results in the Supplement).

### 3.3 Robustness against coil tilt inaccuracies

Overall, for both healthy individuals (Figure 4A) and MDD patients (Figure 4B), the fraction of FPN, CON/AMN, and SN covered by E-field hotspots resulting from the heuristic F5-AF7 TMS coil placement did not significantly deviate from the ideal 0 degree tilt placement for any of the four tested coil tilt conditions (-10, -5, +5, +10 degrees) along both the y- and x-axis of the TMS coil (all Cohen’s |d|’s ≤ 0.35). One exception was seen for SN coverage in the group of healthy individuals: When tilting the TMS coil along the y-axis, the most medial tilt of the TMS coil (i.e., +10 degrees) resulted in significantly more E-field hotspot coverage of the SN in comparison to the ideal 0 degree tilt condition (p_FDR_ < .001, d = 0.50; Figure 4A, upper row, right).

**Figure 4.**
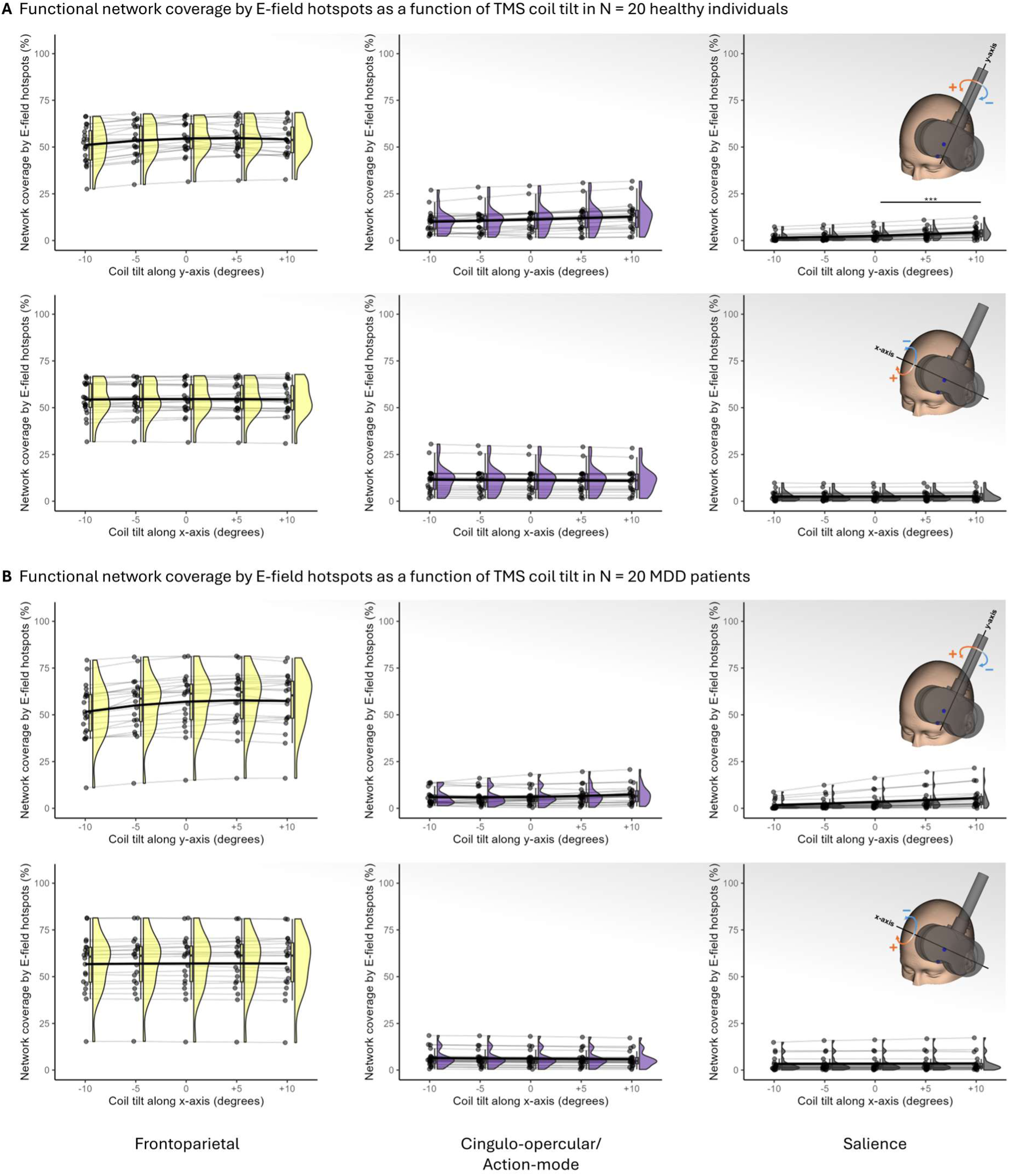
Percent coverage of functional networks (Lynch et al. (31) parcellation) by simulated E-field hotspots as a function of different TMS coil tilts (-10, -5, +5, +10 degrees) along both the y-axis (upper row) and x-axis (lower row) of the TMS coil. Coverage of the frontoparietal network (FPN; left column), cingulo-opercular/action mode network (CON/AMN; middle column), and salience network (SN; right column) by simulated E-field hotspots in (**A**) healthy individuals (N=20) and (**B**) MDD patients (N=20). Note that the depicted values represent the average over ten different percentile-based E-field thresholds (see Methods and Materials for more details). The condition-specific means across all participants are connected by thick black lines. *** p_FDR_ < .001 and d ≥ 0.50.

### 3.4 Importance of TMS coil orientation

As can be seen from Figure 5A and B for healthy individuals and MDD patients, respectively, E-field hotspot coverage of the FPN decreases, while coverage of the CON/AMN and SN increases, with increasing angular deviation from the optimal 0 degree coil orientation. For details of statistical significance and effect sizes for the respective pair-wise comparisons, see Figure S2 in the Supplement.

**Figure 5.**
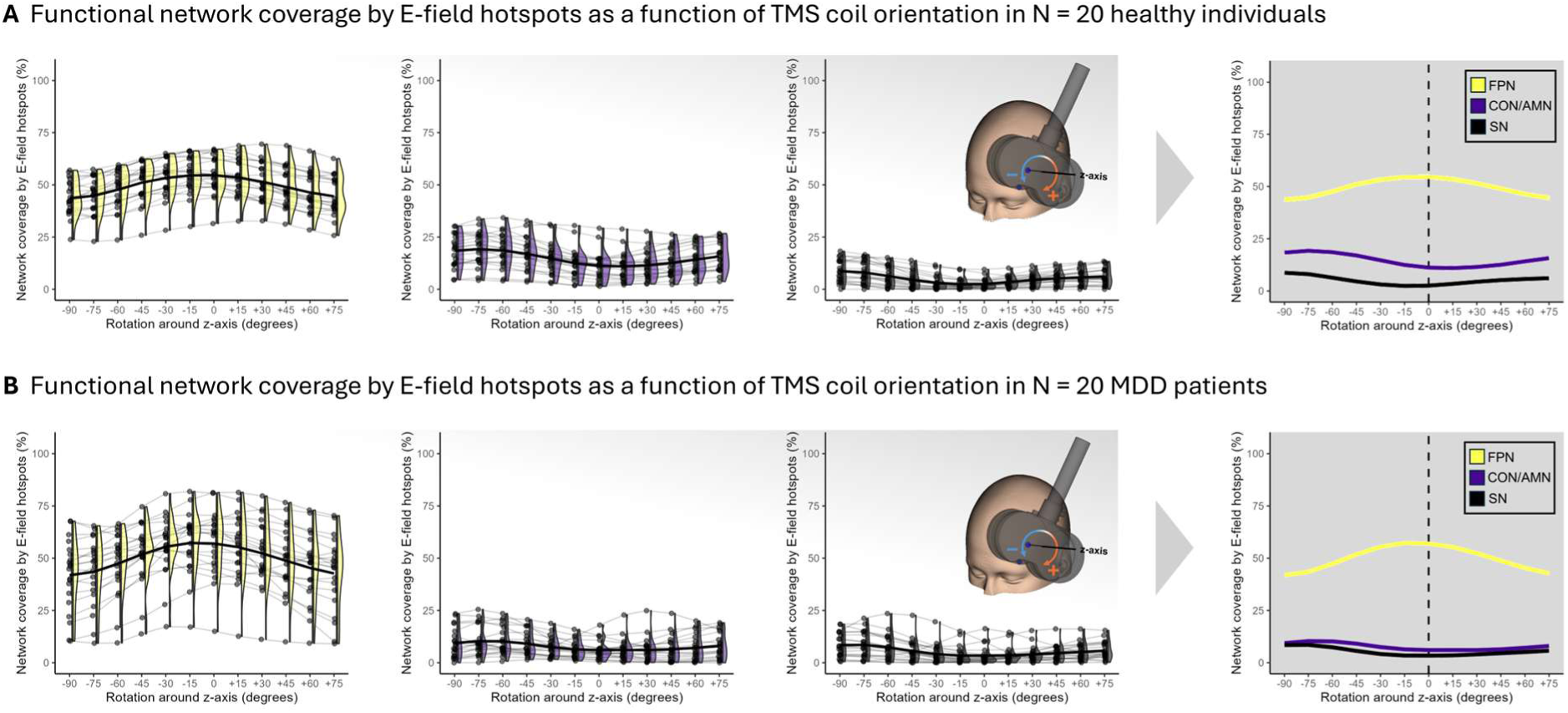
Percent coverage of functional networks (Lynch et al. (31) parcellation) by simulated E-field hotspots as a function of different TMS coil rotations (-90 to +75 degrees, in 15 degree steps) around the z-axis of the TMS coil. Coverage of the frontoparietal network (FPN), cingulo-opercular/action mode network (CON/AMN), and salience network (SN) by simulated E-field hotspots in (**A**) healthy individuals (N=20) and (**B**) MDD patients (N=20). Note that the depicted values represent the average over ten different percentile-based E-field thresholds (see Methods and Materials for more details). The condition-specific means across all participants are connected by thick black lines, which are also summarized for all three functional networks in the panels on the right. For details of statistical significance and effect sizes for the respective pair-wise comparisons, see Figure S2 in the Supplement.

## 4 Discussion

In the present study, we used functional brain network- and E-field-guided optimization of TMS on a population level (i.e., in MNI standard space; (71)) to derive a standard TMS coil placement for reliable stimulation of the depression-relevant frontoparietal network (FPN; Figure 1B). We also provided heuristics to reproduce this coil placement in any given individual based on EEG electrode positions and corresponding SGP-based (57) scalp measurements (Figure 1C). Importantly, these heuristics are easily and immediately applicable in clinical practice, without the need for individual neuroimaging and -navigation.

In a standard brain template and in individual brains of N = 20 healthy participants and N = 20 MDD patients fulfilling the clinical criteria for TMS treatment, in contrast to a standard placement of the TMS coil widely used in depression treatment (i.e., TMS coil centered at the Beam F3 position and oriented 45 degrees relative to the sagittal midline; (26)), the population-based FPN-optimized TMS coil placement and its F5-AF7 EEG electrode-based heuristic (i.e., TMS coil centered at the F5 EEG electrode position with its handle oriented along the F5-AF7 axis) resulted in a considerably larger coverage of the FPN by the hotspots of simulated E-fields (Figure 2, Figure 3, Table 2). This stronger and more selective FPN targeting was (largely) robust against slight inaccuracies in TMS coil placement (i.e., tilts along the y- and x-axis of the TMS coil) that might occur in clinical practice (Figure 4). We additionally showed that the proposed optimal angle of the TMS coil, i.e., parallel to the sagittal midline or along the F5-AF7 axis, is an important factor for selective FPN engagement (Figure 5, Figure S2) and in line with previous findings (9). The overall pattern of results was replicated in two (revised versions of) commonly used functional brain network parcellations (cf. results in the Supplement).

The relevance of the FPN in depression has been shown in numerous studies on functional brain activation and connectivity abnormalities in MDD (e.g., (60,72–75)). Dysfunction of the FPN has often been associated with deficits in cognitive control, emotion regulation, and working memory – symptoms commonly observed in MDD. More importantly, while most previous studies were correlational in nature, a network strongly resembling the FPN has been causally implicated in depression, based on combined evidence from lesion, deep brain stimulation, and TMS data (Siddiqi, Schaper, et al., 2021; Padmanabhan et al., 2019). In addition, grey matter changes after electroconvulsive therapy (76) as well as cortical E-fields induced by H1 TMS coils (77) – both treatments associated with (strong) antidepressant efficacy (78–81) – were also preferentially connected to a network similar to the causal frontoparietal depression circuit. Recent work using concurrent TMS and EEG (TMS-EEG) has furthermore shown that two forms of accelerated rTMS (intermittent and continuous theta-burst stimulation to the left and right DLPFC, respectively) normalize current propagation within and excitability of the FPN, which correlates with antidepressant efficacy (82). The FPN therefore appears to be a highly promising neuromodulation target for TMS depression treatment.

The present study is neither the first to propose a scalp measurement-based approach for targeting the Siddiqi et al. (38) causal depression circuit (cf. (83,58)), nor the first to propose the F5 EEG electrode position as a promising heuristic for guiding TMS coil placement in depression treatment (84,85). However, previous approaches were merely based on simple scalp projections of single voxel coordinates (83,58) or focused on anatomical targeting of the DLPFC (84,85), ignoring the (spatially extended) topography of functional (target) networks across the brain. In contrast, our coil placement (heuristic) has been derived by maximizing the topographic overlap between the target network (cluster) and simulated TMS-induced E-fields. We furthermore not only provide a (heuristic for the) optimal position of the TMS coil center, but also an optimal TMS coil orientation, which is often neglected in other approaches (56), but can considerably influence targeting specificity (i.e., on-vs. off-target stimulation; (7,23)) and functional network engagement (9).

Following the anatomical location of the FPN within the DLPFC, our derived population-based FPN-optimized TMS coil placement and its F5-AF7 EEG electrode-based heuristic are more lateral and anterior than placements currently used in clinical practice. This is in line with findings showing preferential targeting of the FPN (9) and better depression treatment outcomes (e.g., (16,86)) when using more lateral and anterior TMS coil placements. Of relevance, the individualized, sgACC-FC-guided TMS targets (and derived coil placements) of the Stanford Neuromodulation Therapy (SNT) protocol, which has so far resulted in excellent response and remission rates (18,20,21), are also more laterally and anteriorly localized than the standard (Beam) F3 location (cf. Figure 2 in (21), and Figure 1 in (20)). This and other findings have led Wu and Baeken (87) to argue that, in comparison to the classical (dorsolateral) F3 location, TMS applied at more lateral and anterior (i.e., ventrolateral) sites may be generally more effective for depression treatment.

At the same time, the more lateral and anterior FPN-optimized coil location may be more unpleasant and less tolerable for patients (e.g., in terms of peripheral co-stimulation of cranial nerves and muscles; (88)), especially at higher intensities (e.g., the commonly used 120% of the resting motor threshold (RMT); e.g., (89,4)). The trade-off between lower tolerability (more discomfort/pain, muscle twitches, etc.) and potentially higher treatment effectiveness will need to be studied in future clinical trials. Given that tilting the coil along its y- and x-axis appears to have only minor effects on FPN targeting (cf. Figure 4), slightly tilting the coil more medially and/or posteriorly may help to reduce discomfort in the individual case. The issue of potential unpleasantness of more anterolateral prefrontal TMS also provokes considerations about (i) alternative targets, i.e., different entry points into the FPN (e.g., via its node(s) in the parietal cortex; (90)) that allow less unpleasant TMS coil placements; and (ii) dosing, i.e., the use of lower intensities to diminish potential discomfort (e.g., using 90% instead of 120% of the RMT, as done in the SNT protocol; (18,20,21)), which might also reduce co-stimulation of non-target networks and, thus, more selective engagement of the target network (7).

While the FPN is a promising target for TMS treatment of depression, it is most likely not the only network that is affected and should be targeted in depression. In fact, previous work suggests that different networks might be relevant for different symptoms (91). Examining the relationship between the naturally occurring interindividual variability in TMS target sites and improvement in different symptoms of depression, Siddiqi et al. (92) used a data-driven clustering approach to derive two major symptom clusters that responded to stimulation of two differentiable functional networks or circuits: A “dysphoric” cluster included symptoms such as sadness, anhedonia, and suicidality, and was preferentially responsive to the classical dorsolateral prefrontal cortex stimulation. In contrast, an “anxiosomatic” cluster associated with insomnia, irritability, and sexual disinterest responded better to more dorsomedially targeted TMS. This is in line with findings by Drysdale et al. (93) who derived four distinct sub- or biotypes of depression from correlations between clinical and resting-state fMRI-based imaging markers, one of which was defined by greater anxiety and insomnia and responded particularly well to TMS of the dorsomedial PFC. Likewise, Kaster et al. (94) showed that anxiety symptoms were significantly less responsive to TMS of the dorsolateral PFC than, e.g., mood-related symptoms. Following this, dorso- or even ventrolateral stimulation of the FPN might be primarily suited for the treatment of dysphoric (and potentially cognitive) symptoms, while dorsomedial stimulation of a circuit resembling the default mode network (DMN) appears to be more promising for anxiety-related symptoms (92,95–97).

While stronger and more selective TMS of the FPN appears to be a promising avenue for improving the clinical effectiveness of TMS treatments in depression, the present work so far only offers a theoretical recommendation for a TMS coil placement that has been optimized for prefrontal FPN stimulation. Future prospective randomized controlled clinical trials are necessary to test and compare the treatment effectiveness of using our proposed population-based FPN-optimized TMS coil placement and/or its clinical F5-AF7 EEG electrode-based heuristic against other, established and widely used clinical targeting approaches (e.g., the Beam F3 method). The heuristic F5-AF7 approach presented in this paper enables researchers to carry out such clinical trials without the need for individual costly and time-consuming MR imaging, targeting computations, and neuronavigation, thus increasing clinical feasibility and facilitating large sample sizes.

## Data Availability

All data produced in the present study are available upon reasonable request to the authors.

## Acknowledgements

**Til Ole Bergmann** and **Florian Müller-Dahlhaus** have received funding related to this work from the German Research Foundation (DFG Grant No. 525127358).

**Maximilian Lueckel**: Conceptualization, Methodology, Software, Validation, Formal Analysis, Investigation, Data Curation, Writing - Original Draft, Visualization; **Kathrin Kachel**: Investigation, Data Curation, Writing - Review & Editing; **Jan Engelmann**: Writing - Review & Editing; **Til Ole Bergmann**: Resources, Writing - Review & Editing, Supervision, Funding Acquisition; **Florian Müller-Dahlhaus**: Conceptualization, Writing - Review & Editing, Supervision, Funding Acquisition.

We thank Micheal D. Fox, MD, PhD, for providing the NIfTI file of the lesion depression circuit from Padmanabhan et al. (2019). This manuscript has been published as a preprint on medRxiv.

## Disclosures

None of the authors report any financial interests or potential conflicts of interest.

## Notes

### Competing Interest Statement

The authors have declared no competing interest.

### Funding Statement

Til Ole Bergmann and Florian Mueller-Dahlhaus have received funding related to this work from the German Research Foundation (DFG Grant No. 525127358).

### Author Declarations

The Ethics committee of the State Medical Association Rhineland-Palatinate gave ethical approval for this work.

