## Supplement for "A standard coil placement for reliable transcranial magnetic stimulation of the frontoparietal depression network: the ‘F5-AF7 method‘"

Short title:

**TMS targeting of the frontoparietal depression network**

Maximilian Lueckel[1,2,†], Kathrin Kachel[3,2], Jan Engelmann[3,2],

Til Ole Bergmann[2,1,*], Florian Müller-Dahlhaus[3,2,*,†]

[1] Leibniz Institute for Resilience Research (LIR) gGmbH, Mainz, Germany

[2] Neuroimaging Center (NIC), Focus Program Translational Neuroscience (FTN), Johannes Gutenberg University Medical Center, Mainz, Germany

[3] Department of Psychiatry and Psychotherapy, Johannes Gutenberg University Medical Center, Mainz, Germany

* Contributed equally

[†]Corresponding authors:

Maximilian Lueckel:

Florian Müller-Dahlhaus:

#### Methods and Materials

*Sample*

N = 20 healthy participants (15 female, 5 male; age: M = 25.35 years, SD = 3.34 years, range: 20-32 years) without any history of psychiatric or neurological conditions were recruited for a study on concurrent transcranial magnetic stimulation and functional magnetic resonance imaging.

In addition, N = 20 patients with a diagnosis of Major Depressive Disorder (MDD) were recruited as part of the IDeA-L (Individual Determinants of Antidepressant Response) longitudinal study (for details on the study protocol, see preregistration at https://osf.io/q7uxf/files/z8jru). Detailed (clinical) characteristics of the sample can be found in Table S1.

All participants identified as white and were screened by a physician for their eligibility for magnetic resonance imaging and/or transcranial magnetic stimulation. All study procedures were approved by the local ethics committee in accordance with the Declaration of Helsinki (ethics vote no. 2019-14452 and 2020-15333, respectively), and written informed consent was provided by each participant.

**Table S1**. Characteristics of MDD patient sample (N = 20).

| | | |
|---|---|---|
| **Age** (years) | | 42.40 (13.01); 23-64 |
| **Sex** | | |
| | Women | 7 (35%) |
| | Men | 13 (65%) |
| **Duration of education** (years) | | |
| | 10-12 | 2 (10%) |
| | >12 | 18 (90%) |
| **Age of onset** (years) | | 30.65 (13.70); 15-55 |
| **Total number of depressive episodes** | | 4.75 (3.14); 2-11 |
| **IDS score*** | | 33.40 (10.72); 19-52 |
| **MADRS score*** | | 24.60 (7.96); 13-40 |
| **Depressive episode duration** (months)* | | 20.35 (12.25); 2-13 |
| **Receiving pharmacotherapy*** | | 19 (95%) |
| **Failed antidepressants** | | |
| | Two | 13 (65%) |
| | Three | 5 (25%) |
| | More than three | 2 (10%) |
| **Previous treatments** | | |
| | Psychotherapy | 15 (75%) |
| | rTMS | 1 (5%) |
| | Electroconvulsive therapy | 0 (0%) |
| **Comorbidities** | | |
| | Double Depression | 5 (25%) |
| | Generalized Anxiety Disorder | 1 (5%) |
| | Panic Disorder | 2 (10%) |
| | Social Phobia | 2 (10%) |
| | Attention Deficit Hyperactivity Disorder | 1 (5%) |

Note: Numbers represent means or absolute numbers of patients, brackets contain standard deviations or proportions of patients relative to the whole sample (N = 20), respectively. Value ranges are given where appropriate. * indicates information recorded at the time point of the fMRI scan. IDS = Inventory of Depressive Symptoms (1,2); MADRS = Montgomery-Åsberg Depression Rating Scale (3).

*Acquisition and processing of individual subject MRI data*

All MRI data were acquired on a Siemens Magnetom Prisma 3T scanner, using a 32-channel Siemens head coil. Respectively, for healthy individuals and MDD patients, we acquired a single ~15 minutes run (650 volumes) and two ~10 minutes runs (2 x 404 volumes) of resting-state fMRI data, using a T2*-weighted multi-band, multi-echo (MBME) echo-planar imaging (EPI) sequence (repetition time (TR): 1435 ms; echo time $(TE)_{1/2/3/4}$: 12/28.24/44.48/60.72 ms; field-of-view (FOV): 215 mm; flip angle (FA): 65°; 2.5 mm isotropic voxels; 51 slices; anterior-posterior (AP) phase encoding direction; in-plane acceleration factor: 2; multi-band acceleration factor: 3). During resting-state, participants were instructed to stay awake, keep their eyes open (while looking at a fixation cross), and let their mind wander without focusing on a particular thought. In addition, high-resolution (MPRAGE) T1-weighted (T1w; TR: 2500 ms; TE: 2.22 ms; FOV: 256 mm; FA: 8°) and T2-weighted (T2w; TR: 3200 ms; TE: 563 ms; FOV: 256 mm) anatomical images (0.8 mm isotropic voxels; 208 sagittal slices) were acquired.

After acquisition, all anatomical and functional MRI data were converted to the brain imaging data structure (BIDS) format (4). Before regular preprocessing, anatomical (T1w and T2w) images were subjected to FastSurfer for accelerated brain surface reconstruction (5), while NORDIC denoising was applied to each functional run and echo individually, to reduce thermal noise in the data (using only the magnitude images and a theoretical thermal noise level) (6,7). We then used fMRIPrep (version 25.1.1; (8)) for minimal preprocessing of the data, incorporating the brain surfaces already generated by FastSurfer. In short, an echo-planar imaging (EPI) reference volume was generated from the shortest echo of each functional run, using a custom methodology of fMRIPrep. Fieldmaps were estimated for each run based on two corresponding EPI references with opposing phase-encoding directions using topup (9) and aligned with rigid-registration to the EPI reference volume. Afterwards, motion correction was performed using mcflirt from FSL (10), realigning all volumes to the EPI reference volume. In addition, slice time correction was performed using 3dTShift from AFNI (11), realigning all slices in time to the middle of each TR. The EPI reference volume was then co-registered to the T1w reference using mri_coreg from FreeSurfer (12), followed by FSL's flirt (13) with the boundary-based registration (14) cost-function and six degrees of freedom. Functional time series were further projected to the individually reconstructed cortical surface and resampled to left-/right-symmetric "fsLR" space using Connectome Workbench utilities (15), excluding voxel time series with a locally high coefficient of variation. All resamplings were performed with a single interpolation step by composing all respective transformations (i.e., head-motion transform matrices, susceptibility distortion correction, and co-registrations to the anatomical space).

The minimally preprocessed functional echoes (in volume space) were then subjected to the tedana workflow (version 25.0.1) for TE-dependence analysis (16). More specifically, a monoexponential model was fit to the data at each voxel using nonlinear model fitting in order to estimate T2* and S0 maps. Afterwards, the multi-echo data were optimally combined using the T2* combination method (17). Principal component analysis (PCA) was applied to the optimally combined data for dimensionality reduction, before independent component analysis (ICA) was used to decompose the dimensionally reduced dataset. Kappa and Rho values were calculated as measures of TE-dependence and TE-independence, respectively. Next, component selection was performed to automatically identify BOLD (TE-dependent) and non-BOLD (TE-independent) components using the tedana_orig decision tree (which is very similar to the criteria of the MEICA v2.5 decision tree by Kundu et al. (18); https://tedana.readthedocs.io/en/stable/included_decision_trees.html). Despite tedana's automatic classification into noise and signal components, all resulting ICA components were visually reviewed and discarded as noise (or accepted as signal of interest) if necessary, following the criteria detailed in (19,20). Time series of rejected components were then orthogonalized with respect to those of accepted components.

The eXtensible Connectivity Pipeline-DCAN (XCP-D; version 0.11.0; (21–23)) was used to post-process the outputs of fMRIPrep and tedana. More specifically, native-space T1w images were transformed to MNI152NLin2009cAsym space at 1 $mm^3$ resolution. In addition, HCP-style midthickness, inflated, and very-inflated surfaces were generated from the white-matter and pial fsLR-space surface meshes (15) and transformed to align with the volumetric template. Framewise displacement (FD) was calculated from the low-pass filtered motion parameters (24,25) and volumes with FD > 0.3 mm were flagged as high-motion outliers for later censoring (25). Additionally, the BOLD data were despiked with AFNI 's 3dDespike. Nuisance regressors included tedana-derived, orthogonalized noise IC timecourses, the mean grey matter (i.e. global), white matter, and CSF signal, the six motion parameters, their temporal derivative, their quadratic expansion, as well as the temporal derivative of their quadratic expansion. The (surface-projected) time series and confounds were band-pass filtered using a second-order Butterworth filter, in order to retain signals between 0.01-0.1 Hz. The resulting time series were then denoised via linear regression. Lastly, the denoised BOLD data was smoothed using Connectome Workbench with a Gaussian kernel (FWHM = 6.0 mm) and, if multiple runs were available, concatenated after standardizing the time series.

### Results

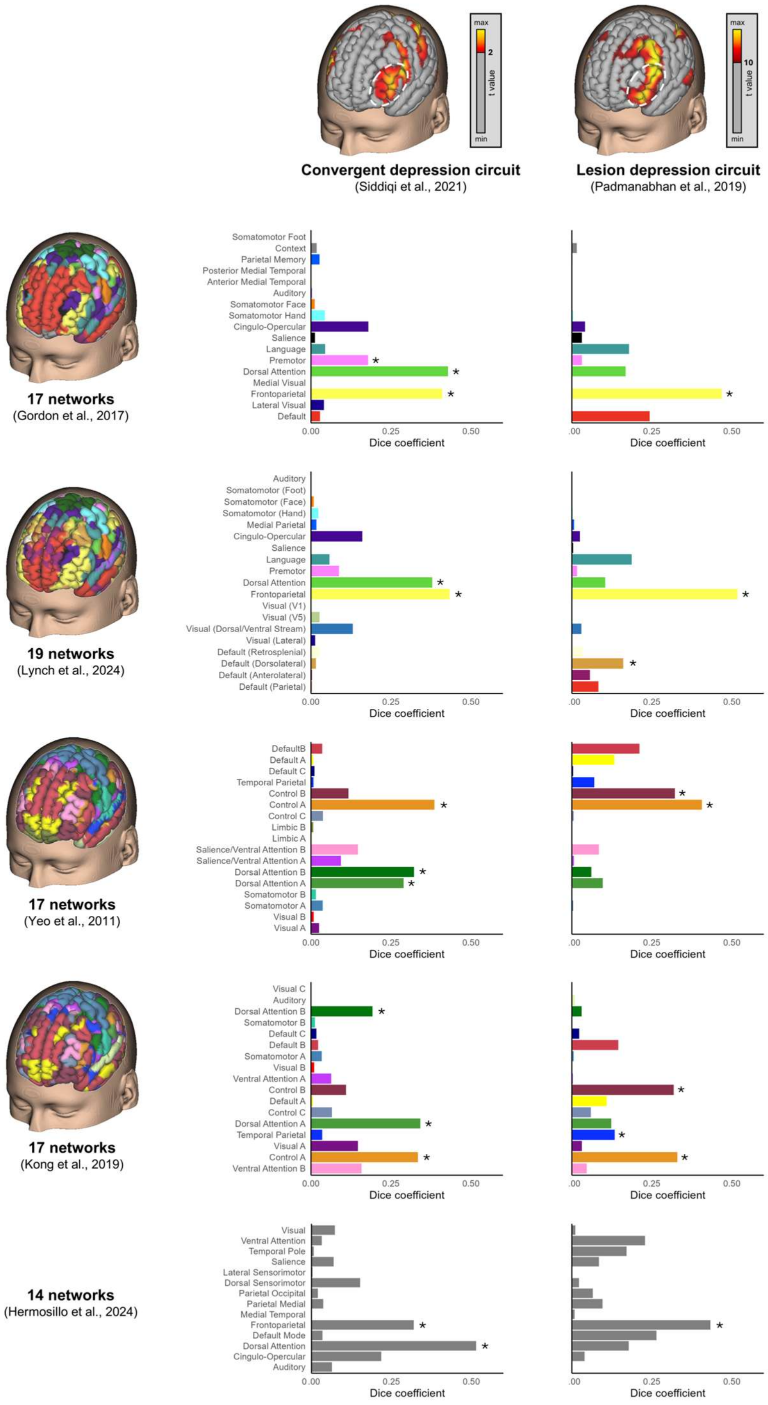


**Figure S1.** Bar plots of Dice coefficients and their statistical significance (*p < .05) for the spatial overlap between (surface-mapped and thresholded versions of) the two depression circuit maps by Siddiqi et al. (26) and Padmanabhan et al. (27) and functional network parcellations by i) Gordon et al. (28) (17 networks), ii) Lynch et al. (29) (a refined version of the Gordon parcellation with 19 networks), iii) Yeo et al. (30) (17 networks), and iv) Kong et al. (31) (a refined version of the Yeo parcellation with 17 networks), as well as v) probabilistic maps of 14 commonly observed large-scale functional networks, provided as part of a precision functional brain atlas by Hermosillo et al. (32).

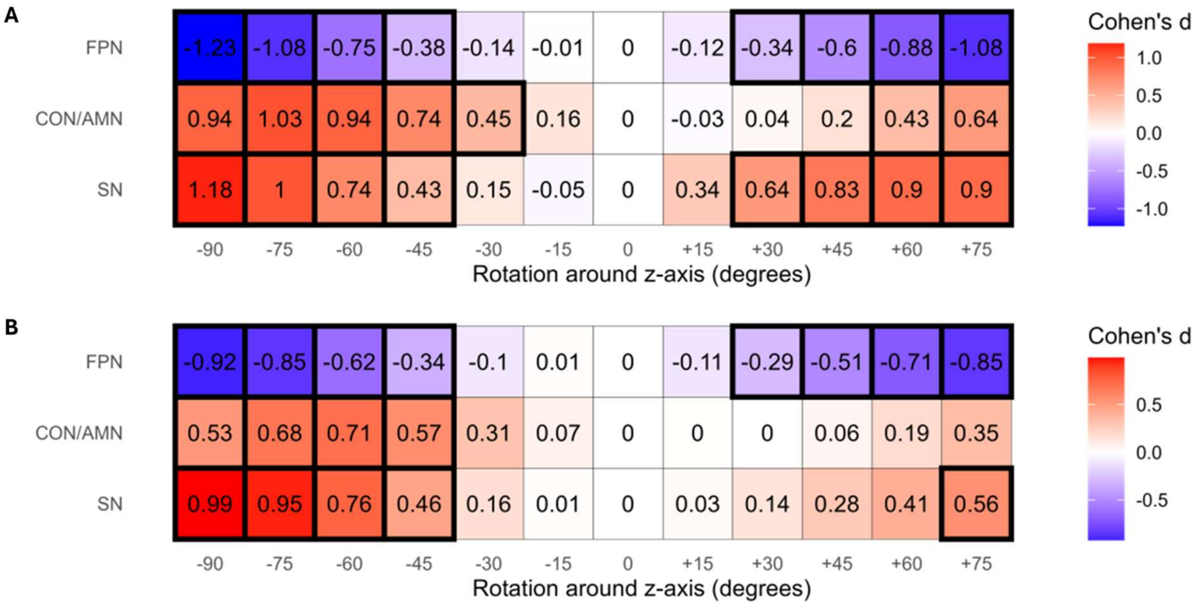


**Figure S2.** Effect size and statistical significance for changes in coverage of the frontoparietal network (FPN; top row), cingulo-opercular/action mode network (CON/AMN; middle row), and salience network (SN; bottom row) by simulated E-field hotspots as a function of different TMS coil rotations (-90 to +75 degrees, in 15 degree steps) around the z-axis of the TMS coil, compared against the original heuristic F5-AF7 EEG electrode-based TMS coil placement (with 0 degrees rotation), in (**A**) healthy individuals (N=20) and (**B**) MDD patients (N=20). Effects sizes (Cohen's d) are color-coded, while statistically significant effects (with an FDR-corrected $p < .05$) are marked by a thick black outline.

*Validation of* the population-based *FPN-optimized and heuristic F5-AF7 TMS coil placement using the functional network parcellation by Kong et al.* (31)

Figure S3 illustrates and Table S2 summarizes the (average) percent coverage of the Control A and B network (equivalent to the FPN of the Lynch/Gordon parcellations), Ventral Attention A and B network (equivalent to the CON/AMN of the Lynch/Gordon parcellations), and Control C network (equivalent to the SN of the Lynch/Gordon parcellations) by simulated E-field hotspots resulting from a standard clinical TMS coil placement (i.e., TMS coil centered on the Beam F3 EEG electrode position and oriented 45 degrees relative to the sagittal midline), the population-based FPN-optimized TMS coil placement, and its F5-AF7 EEG electrode-based clinical heuristic (i.e., TMS coil centered on the F5 EEG electrode position with its handle oriented along the F5-AF7 axis) in MNI standard space, healthy individuals (N=20), and MDD patients (N=20).

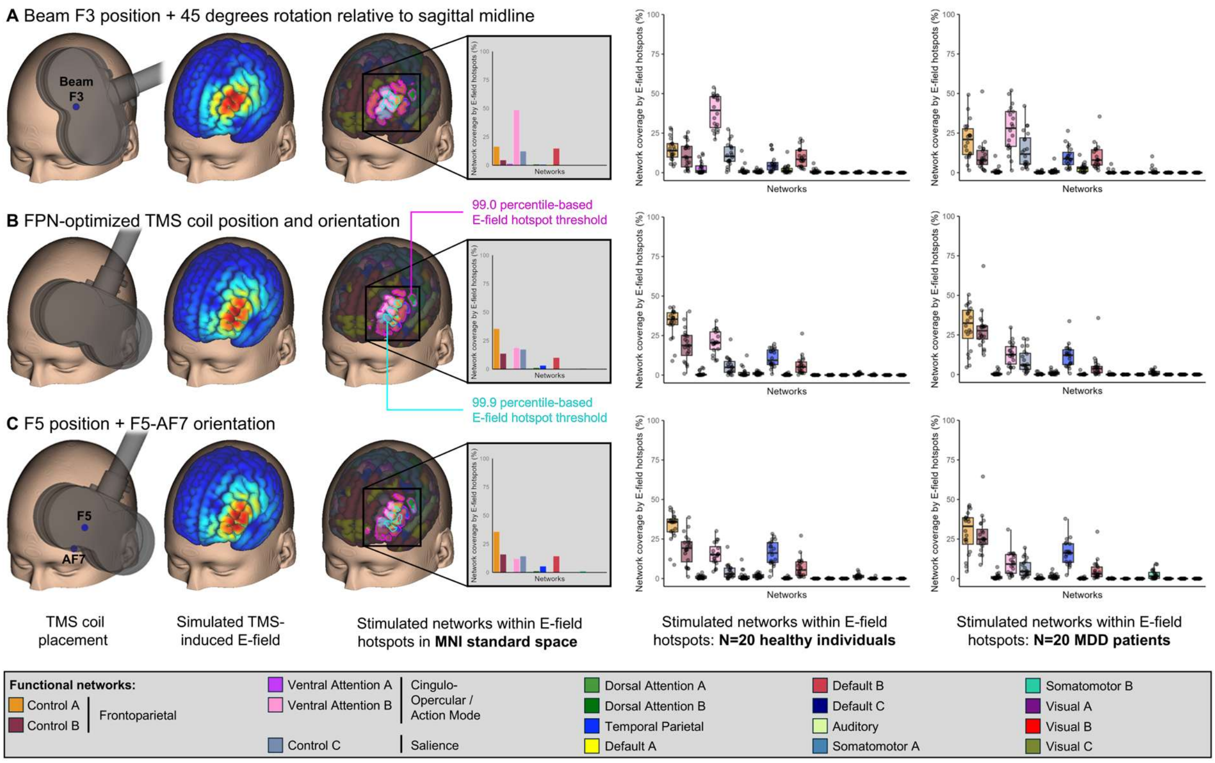


**Figure S3. Percent coverage of functional networks (Kong et al.** (31) **parcellation) by simulated E-field hotspots.** Functional network coverage resulting from (**A**) a standard clinical TMS coil placement (i.e., TMS coil centered on the Beam F3 position and oriented 45 degrees relative to the sagittal midline), (**B**) the population-based FPN-optimized TMS coil placement, and (**C**) its F5-AF7 EEG electrode-based heuristic (i.e., TMS coil centered on the F5 EEG electrode position with its handle oriented along the F5-AF7 axis) in MNI standard space, healthy individuals (N=20), and MDD patients (N=20). Note that the depicted values represent the average over ten different percentile-based E-field thresholds (see Methods and Materials for more details). Exemplary E-field hotspots are illustrated for the lowest (99.0 percentile; magenta outline) and highest (99.9 percentile; cyan outline) threshold.

**Table S2**. Functional network coverage by E-field hotspots of different TMS coil placements.

| TMS coil placement | Network coverage by E-field hotspots (%) | | | | | | | | | | | | | | | | | | | | | | | | |
|---|---|---|---|---|---|---|---|---|---|---|---|---|---|---|---|---|---|---|---|---|---|---|---|---|---|
| | Frontoparietal | | | | | | | | | | Cingulo-opercular/Action-mode | | | | | | | | | | Salience | | | | |
| | Control A | | | | | Control B | | | | | Ventral Attention A | | | | | Ventral Attention B | | | | | Control C | | | | |
| | MNI | HC | | MDD | | MNI | HC | | MDD | | MNI | HC | | MDD | | MNI | HC | | MDD | | MNI | HC | | MDD | |
| | | M | SD | M | SD | | M | SD | M | SD | | M | SD | M | SD | | M | SD | M | SD | | M | SD | M | SD |
| Beam F3 (+45 deg) | 16.37 | 14.62 | 6.99 | 20.65 | 12.10 | 4.57 | 11.20 | 8.09 | 10.67 | 10.90 | 1.33 | 3.14 | 4.02 | 1.12 | 2.43 | 48.59 | 38.35 | 10.67 | 27.47 | 14.31 | 12.30 | 11.53 | 7.22 | 14.47 | 11.43 |
| FPN-optimized | 35.34 | 32.88 | 9.31 | 30.00 | 13.97 | 13.56 | 19.07 | 10.23 | 28.19 | 11.94 | 0.0 | 0.91 | 1.53 | 0.71 | 1.28 | 18.78 | 20.92 | 7.22 | 12.79 | 7.33 | 17.25 | 5.67 | 5.44 | 8.64 | 7.17 |
| F5-AF7 | 35.75 | 32.74 | 9.32 | 29.32 | 13.07 | 15.80 | 18.54 | 9.81 | 27.13 | 11.75 | 0.0 | 0.89 | 1.05 | 1.17 | 1.77 | 12.31 | 15.54 | 6.97 | 9.77 | 7.42 | 14.34 | 4.33 | 4.71 | 6.43 | 5.59 |

Note: E-field hotspot coverage of other networks than the Control A, Control B, Ventral Attention A, Ventral Attention B, and Control C networks is not explicitly reported here, but see Figure S3.

Statistical comparisons between the three targeting approaches showed that, in comparison to the standard Beam F3-based TMS coil placement, E-field hotspots of the newly derived population-based FPN-optimized and heuristic F5-AF7 TMS coil placements covered (i) a significantly larger fraction of the Control A network in both healthy individuals ($p_{FDR} < .001$, $d = 2.18$ and $p_{FDR} < .001$, $d = 2.17$, respectively; Figure S4A) and MDD patients ($p_{FDR} < .001$, $d = 0.70$ and $p_{FDR} < .05$, $d = 0.69$, respectively; Figure S4B), (ii) a significantly larger fraction of the Control B network in both healthy individuals ($p_{FDR} < .001$, $d = 0.81$ and $p_{FDR} < .001$, $d = 0.79$, respectively; Figure S4A) and MDD patients ($p_{FDR} < .001$, $d = 1.52$ and $p_{FDR} < .001$, $d = 1.45$, respectively; Figure S4B), (iii) a significantly smaller fraction of the Ventral Attention B network in both healthy individuals ($p_{FDR} < .001$, $d = -1.86$ and $p_{FDR} < .001$, $d = -2.48$, respectively; Figure S4A) and MDD patients ($p_{FDR} < .001$, $d = -1.08$ and $p_{FDR} < .001$, $d = -1.40$, respectively; Figure S4B), and (iv) a significantly smaller fraction of the Control C network in both healthy individuals ($p_{FDR} < .001$, $d = -0.90$ and $p_{FDR} < .001$, $d = -1.13$, respectively; Figure S4A) and MDD patients ($p_{FDR} < .05$, $d = -0.53$ and $p_{FDR} < .001$, $d = -0.71$, respectively; Figure S4B). For the Ventral Attention A network, only the heuristic F5-AF7 TMS coil placement resulted in a significantly smaller fraction of coverage only in healthy individuals ($p_{FDR} < .05$, $d = -0.53$; Figure S4A).

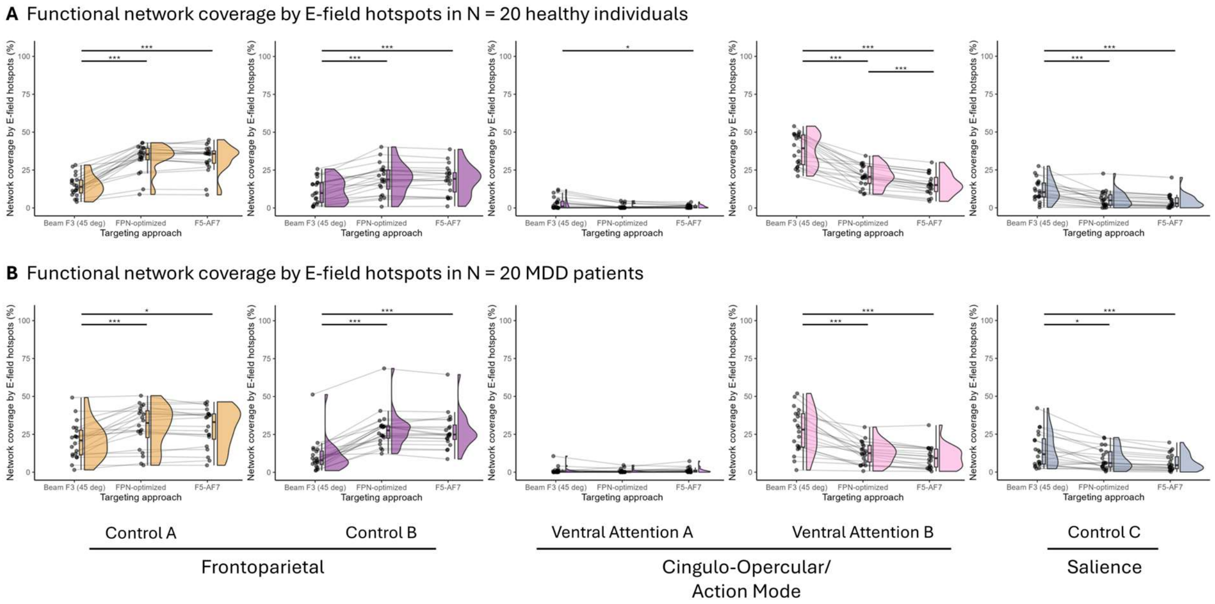


**Figure S4. Percent coverage of functional networks (Kong et al.** (31) **parcellation) by simulated E-field hotspots.** Coverage of the Control A, Control B, Ventral Attention A, Ventral Attention B, and Control C networks by simulated E-field hotspots resulting from a standard clinical TMS coil placement (i.e., TMS coil centered on the Beam F3 position and oriented 45 degrees relative to the sagittal midline), the population-based FPN-optimized TMS coil placement, and its F5-AF7 EEG electrode-based heuristic (i.e., TMS coil centered on the F5 EEG electrode position with its handle oriented along the F5-AF7 axis) in (**A**) healthy individuals (N=20) and (**B**) MDD patients (N=20). Note that the depicted values represent the average over ten different percentile-based E-field thresholds (see Methods and Materials for more details). *** $p_{FDR} < .001$ and $d \geq 0.50$. * $p_{FDR} < .05$ and $d \geq 0.50$.

*Robustness against placement inaccuracies using the functional network parcellation by Kong et al.* (31)

Overall, for both healthy individuals (Figure S5A) and MDD patients (Figure S5B), the fraction of Control A, Control B, Ventral Attention A, Ventral Attention B, and Control C network covered by E-field hotspots resulting from the heuristic F5-AF7 TMS coil placement did not significantly deviate from the ideal 0 degree tilt placement for any of the four tested coil tilt conditions (-10, -5, +5, +10 degrees) along both the y- and x-axis of the TMS coil (all Cohen's |d|'s ≤ 0.43). For the group of healthy individuals, one exception was seen for Ventral Attention A network coverage: When tilting the TMS coil along the y-axis, the most lateral tilt of the TMS coil (i.e., -10 degrees) resulted in significantly more E-field hotspot coverage of the Ventral Attention A network in comparison to the ideal 0 degree tilt condition ($p_{FDR} < .001$, d = 0.62; Figure S5A, upper row). For the group of MDD patients, one exception was seen for Control B network coverage: When tilting the TMS coil along the y-axis, the most lateral tilt of the TMS coil (i.e., -10 degrees) resulted in significantly less E-field hotspot coverage of the Ventral Attention A network in comparison to the ideal 0 degree tilt condition ($p_{FDR} < .05$, d = -0.62; Figure S5B, upper row).

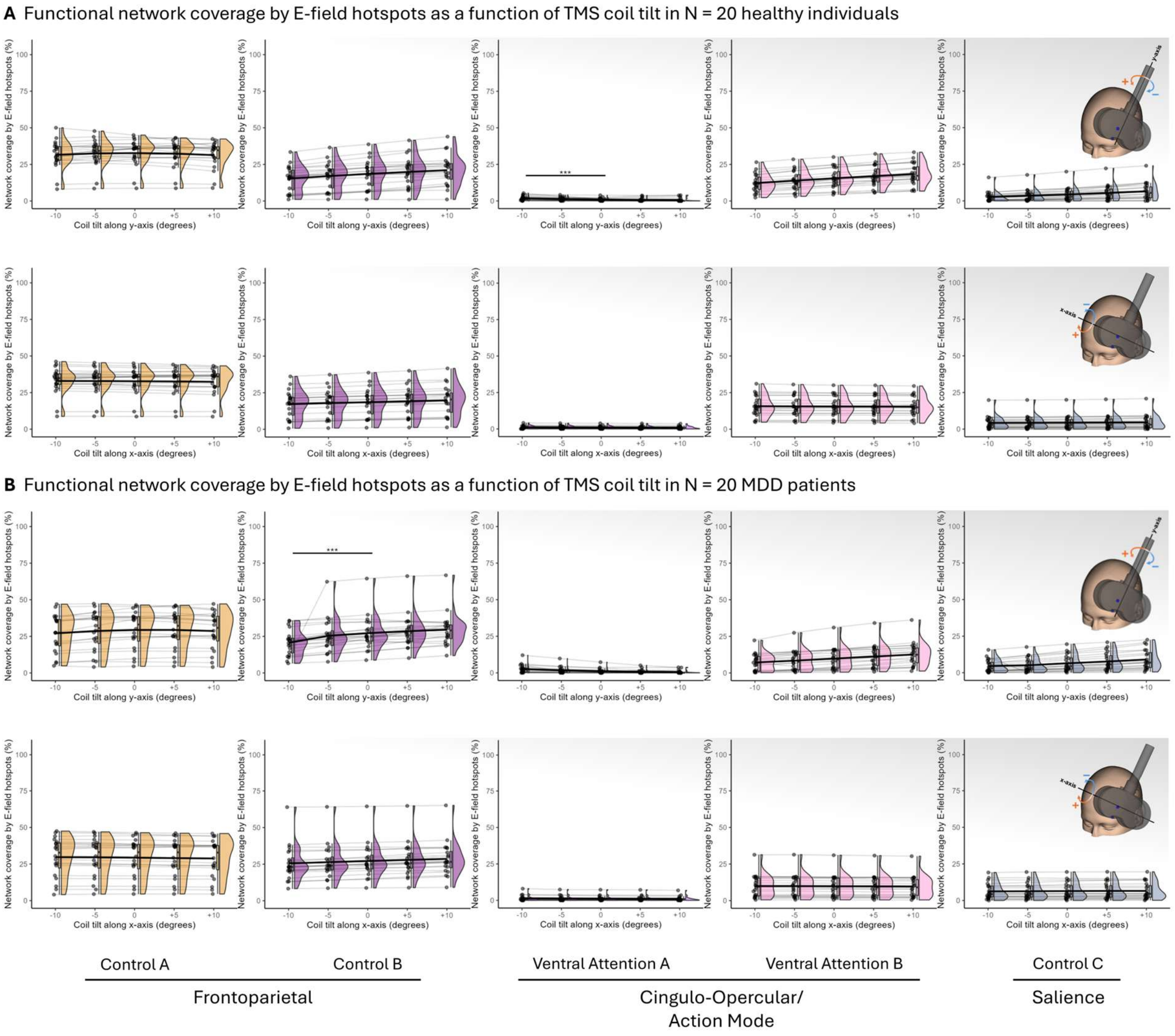


**Figure S5. Percent coverage of functional networks (Kong et al.** (31) **parcellation) by simulated E-field hotspots as a function of different TMS coil tilts (-10, -5, +5, +10 degrees) along both the y-axis (upper row) and x-axis (lower row) of the TMS coil.** Coverage of the Control A, Control B, Ventral Attention A, Ventral Attention B, and Control C networks by simulated E-field hotspots in (**A**) healthy individuals (N=20) and (**B**) MDD patients (N=20). Note that the depicted values represent the average over ten different percentile-based E-field thresholds (see Methods and Materials for more details). *** $p_{FDR} < .001$ and $d \geq 0.50$.

*Importance of TMS coil orientation using the functional network parcellation by Kong et al.* (31)

As can be seen from Figure S6A and B for healthy individuals and MDD patients, respectively, E-field hotspot coverage of the Control A and Control B networks tends to decrease, while coverage of the Ventral Attention A, Ventral Attention B, and Control C networks tends to increase, with increasing angular deviation from the optimal 0 degree coil orientation. For details of statistical significance and effect sizes for the respective pair-wise comparisons, see Figure S7.

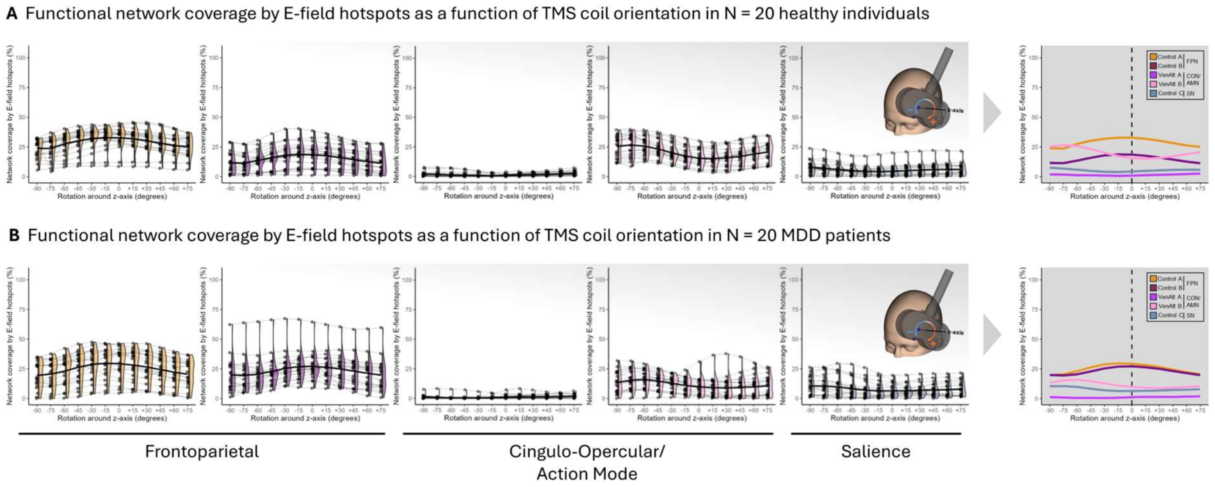


**Figure S6. Percent coverage of functional networks (Kong et al.** (31) **parcellation) by simulated E-field hotspots as a function of different TMS coil rotations (-90 to +75 degrees, in 15 degree steps) around the z-axis of the TMS coil.** Coverage of the Control A, Control B, Ventral Attention A, Ventral Attention B, and Control C networks by simulated E-field hotspots in (**A**) healthy individuals (N=20) and (**B**) MDD patients (N=20). Note that the depicted values represent the average over ten different percentile-based E-field thresholds (see Methods and Materials for more details). The condition-specific means across all participants are connected by thick black lines, which are also summarized for all five functional networks in the panels on the right. For details of statistical significance and effect sizes for the respective pair-wise comparisons, see Figure S7.

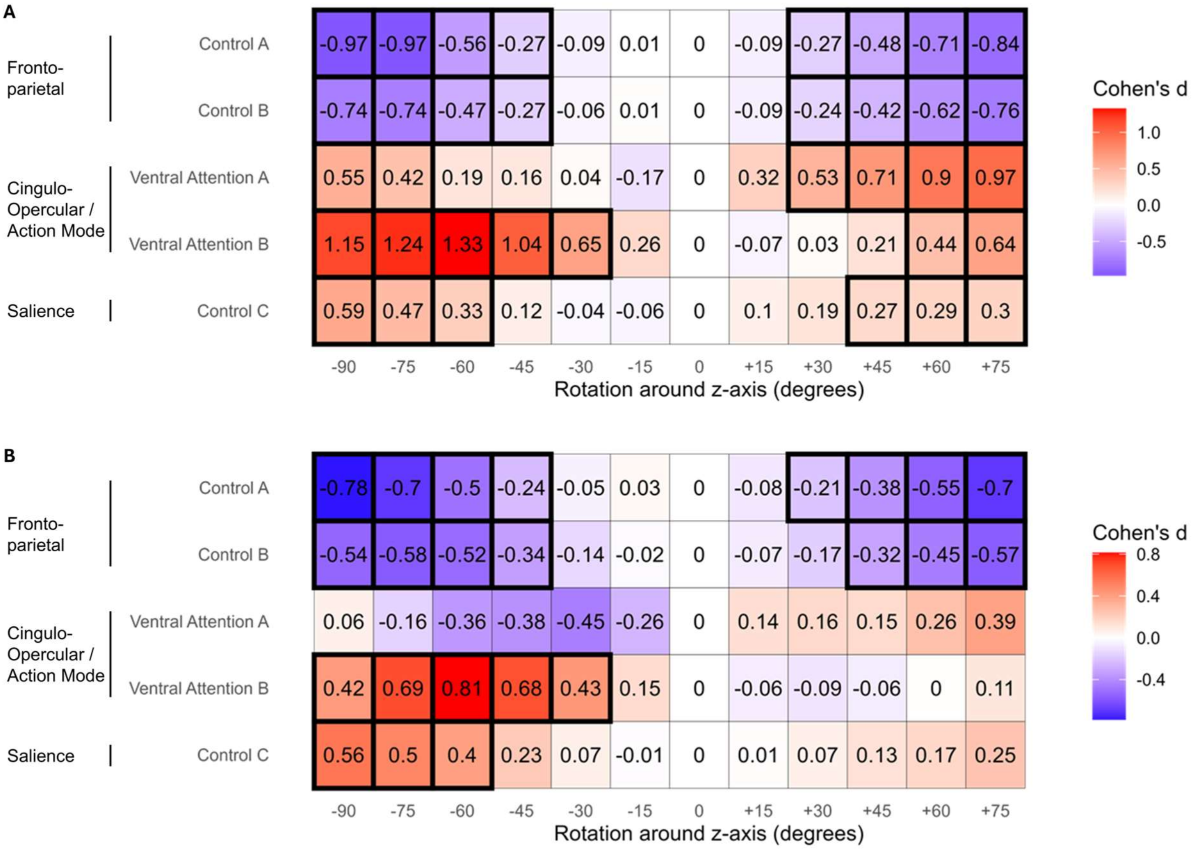


**Figure S7.** Effect size and statistical significance for changes in coverage of the Control A, Control B, Ventral Attention A, Ventral Attention B, and Control C networks by simulated E-field hotspots as a function of different TMS coil rotations (-90 to +75 degrees, in 15 degree steps) around the z-axis of the TMS coil, compared against the original heuristic F5-AF7 EEG electrode-based TMS coil placement (with 0 degrees rotation), in (**A**) healthy individuals (N=20) and (**B**) MDD patients (N=20). Effects sizes (Cohen‘s d) are color-coded, while statistically significant effects (with an FDR-corrected p < .05) are marked by a thick black outline.
